# Country specific mutational profile of SARS-CoV-2 in pre- and post-international travel ban: Effect on vaccine efficacy

**DOI:** 10.1101/2021.02.08.21251359

**Authors:** Sayantan Laha, Raghunath Chatterjee

## Abstract

In order to curb the rapid transmission of SARS-CoV-2, nation-wide lockdowns were implemented as a preliminary measure. Since most countries enforced travel-bans during end of March 2020, the country-specific patterns should be discernible in the subsequent months. We identified frequently mutated non-synonymous mutations in ∼2,15,000 SARS-CoV-2 sequences during pre- and post-travel-ban periods in 35 countries. We further investigated the mutational profile on a bi-monthly basis and traced the progress over the time. Several new mutations have emerged post-travel-ban and on the rise in specific countries, chief among them being A222V and S477N in Spike, and A220V in Nucleocapsid protein. Consequently, we examined the Spike protein epitopes to inspect whether any of these country-specific mutations overlapped with these epitopes. Several mutations were found to be contained within one or more epitopes, including the highly mutated residues of Spike protein, advocating the requirement of active monitoring of vaccine efficacies in respective countries.

## 1. INTRODUCTION

The year 2019 witnessed the emergence of a novel coronavirus, officially designated as SARS-COV-2 by the International Committee on Taxonomy of Viruses (ICTV). SARS-CoV-2 has a single-stranded RNA genome of ∼29kb in length (Kim, Chung et al. 2020). Characterized by the ability to gain rapid mutations in its RNA genome, as commonly seen in RNA viruses (Duffy 2018), constant evolution towards attaining higher stability is often chaperoned by increasing transmissibility and higher virulence. To minimize the transmission of this virus across the globe, country-wide lockdowns had been imposed, towards the end of March 2020. China had been the first to implement travel restrictions across country and state borders as early as January of 2020 (AlTakarli 2020, Cyranoski 2020), while cross-country travel bans were steadily enforced in other countries in the time period ranging from late March to early April (https://www.bbc.com/news/world-52103747). As the number of SARS-CoV-2 isolates were sequenced and made publicly available, one can trace the evolution of the virus through analysing its mutational landscape. Several non-synonymous mutations have been characterized in the early months of the pandemic, chief among which are the substitutions D614G of the Spike glycoprotein and R203K and G204R of the viral Nucleocapsid protein (Laha, Chakraborty et al. 2020). The newly evolved Spike protein variant has been shown to exhibit higher rates of infection, replication and stability (Hou, Wang et al. 2007, Korber, Fischer et al. 2020). These variant strains could be seen to be prevailing across most countries which were a reflection of the inter-country migrations that existed before lockdowns were enforced. It is only in the months following the imposition of the travel restrictions that mutations specific to a particular geographical location, if any should be observed.

Since there is a dearth of medication and therapeutic agents available to combat the SARS-CoV-2 pathogen, engineering vaccines and drugs against this virus has gained top priority(Krammer 2020). Several strategies of vaccine development have been implemented, which are classified as nucleic acid vaccines like DNA and mRNA vaccines, vaccines employing dead, inactivated or attenuated form of the virus, vaccines constructed within carrier vectors and others exemplified by using protein subunits and virus-like particles(Dai and Gao 2020, Dong, Dai et al. 2020). Among the forthcoming protein-based vaccines include PicoVacc, developed by Sinovac,(Dai, Zheng et al. 2020, Zhang, Zeng et al. 2020), BBIBP-CorV(Wang, Zhang et al. 2020, Xia, Zhang et al. 2021), which have successfully been tested in various animal models. Studies have demonstrated that SARS-CoV-2 structural and non-structural proteins can serve as potential candidates for vaccine development, by producing adequate immune responses(Corbett, Edwards et al. 2020, Dhama, Sharun et al. 2020, Dong, Dai et al. 2020).Spike protein is the popular target for epitope constructs specifically due to its ability to elicit strong immune responses within host cells(Cun, Li et al. 2020, Dai, Zheng et al. 2020, Grant, Montgomery et al. 2020, Lan, Ge et al. 2020, Tostanoski, Wegmann et al. 2020, Walsh, Frenck et al. 2020, Wec, Wrapp et al. 2020, Yang, Wang et al. 2020, Yu, Tostanoski et al. 2020, Zhang, Zeng et al. 2020). The engineered variants, termed S-2P has been used as candidate vaccines in various gene-based approaches including mRNA vaccines by Moderna and BioNTech/Pfizer(Corbett, Edwards et al. 2020, Corbett, Flynn et al. 2020, Jackson, Anderson et al. 2020), and the AD26 vaccines developed by Janssen Pharmaceuticals(Mercado, Zahn et al. 2020, Tostanoski, Wegmann et al. 2020). Aside from these, other modifications for further stability enhancement of the Spike protein have been contrived in designing protein vaccines by Novavax(Bangaru, Ozorowski et al. 2020, Tian, Patel et al. 2020). These provided assuring results when tested in animal models and are currently underway through Phase III of clinical trials.

In this study, we compared the mutational profiles of 35 selected countries before and after international travel bans. We have used ∼2,20,000 SARS-CoV-2 sequences and segregated them into two categories based on the time of sequencing; before and after international travel ban. We analysed for substitutions in amino acid sequences throughout all viral proteins. Upon comparing the mutational topography of the sequences in these two phases, we identified the mutations specific to certain countries in the aftermath of the international travel ban. We assessed the temporal variation of these variants over the course of the year, to gain insight into the robustness of these variants. To shed light into the plausible effectiveness of the vaccines, country specific frequently mutated residues were subsequently mapped to the predicted epitopes targeted towards the Spike protein for potential vaccine development.

## 2. MATERIALS AND METHODS

### 2.1 SARS-Cov-2 sequences

A total of ∼2,20,000 SARS-CoV-2 sequences were downloaded from GISAID (https://www.epicov.org/epi3/), available as of November 28^th^ 2020. For each of the ORFs/proteins, partial sequences were filtered out, which accounted for less than 1% of the total isolates. The protein sequences were individually aligned using MUSCLE. The SARS-CoV-2 strain, originating in Wuhan, China (GenBank accession ID **<u>‘NC_045512’</u>**) was considered as the reference strain. Since countries started implementing international travel ban from the third week of March extending up to early April, the isolates were segregated into two groups, based on whether they were sequenced before May (Group B for ‘*before*’) or after May (Group A for ‘*after*’) 2020. We were left with a total of ∼2,15,000 sequences, out of which 1,35,000 sequences are in Group A and 80,000 sequences are in Group B.

### 2.2 Selection of countries for carrying out mutational profiling

We have selected those countries for which there were a minimum of 100 complete sequences, from May to November 28th, provided that these countries also contained a minimum of 30 sequences in the span of January to April. By this measure, we obtained a total of 35 countries in which the mutational characterization was carried out, arranged according to their geographical locations (**Figure 1**)

**Figure 1:**
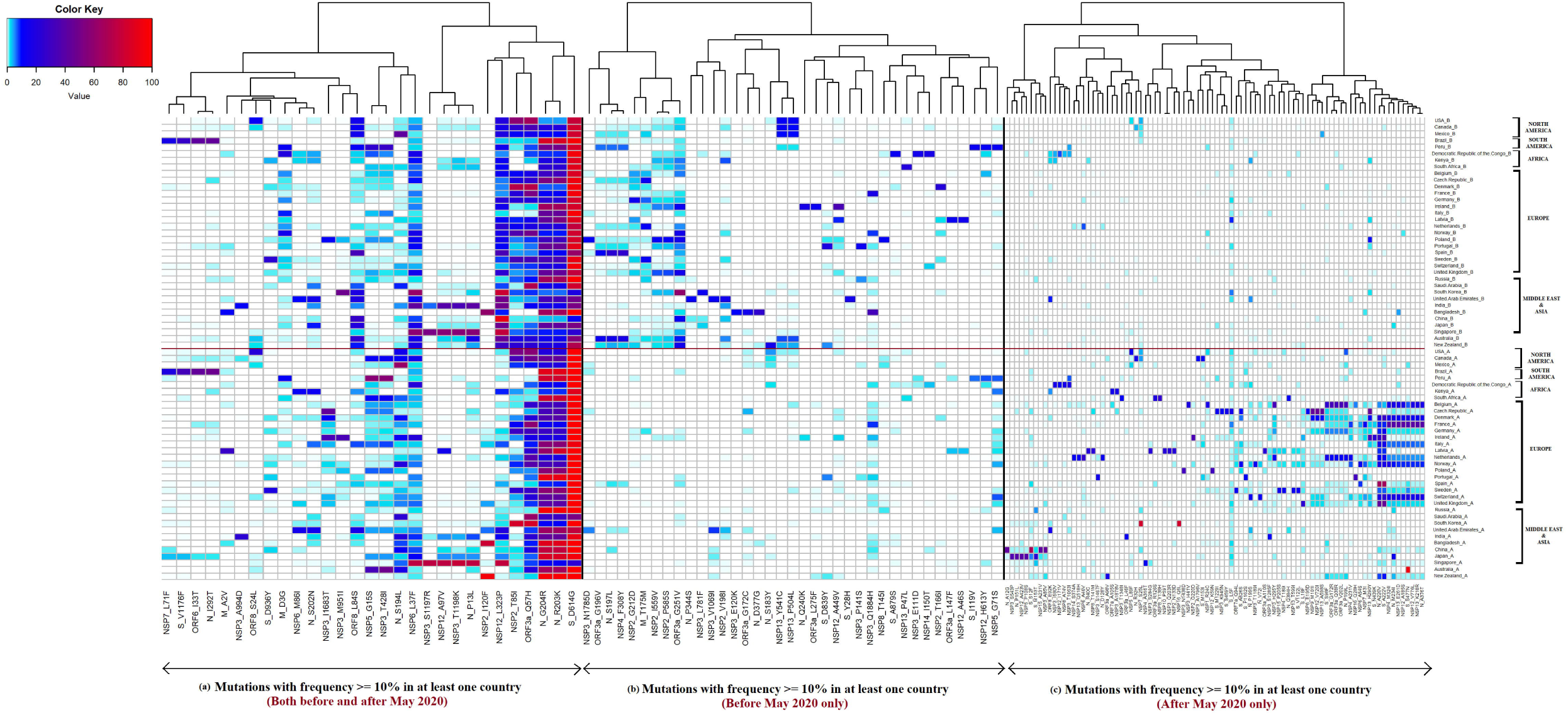
Heat map representing the amino acids substitutions in all SAS-CoV-2 proteins, observed with a frequency of 10% or higher in at least one country in **(a)** both Group A and Group B; **(b)** Group B, and **(c)** Group A only.

### 2.3 Identification of frequently mutated residues for each of the viral proteins

Non-synonymous mutations were identified independently for each of the selected countries for Group A and B. A complete summary with all the unique variants, each characterized by mutations in one or more amino acid positions, along with the frequency of occurrences are calculated. We have defined a substitutions having at least 2% occurrence in all the variants as a frequently mutated residue. An overall summary table depicting the frequencies for all the 35 selected countries, covering each of the identified frequently mutated residues are provided separately for the two groups (**Supplementary Tables1a and 1b**).

### 2.4 Comparison of non-synonymous mutation in pre- and post-international travel ban

In order to obtain a bird’s eye view of the impact of the international travel ban in shaping the mutations in SARS-CoV-2, we have calculated a weighted average of the number of frequently mutated residues for each structural and non-structural protein in 35 selected countries (**Table 1**). The prevailing global trends with the most mutable proteins in both the groups were compared. For the individual countries, both before and after the travel ban were impinged, the number of frequent mutations per 100 amino acids were calculated (**Supplementary Table 2**). The variations in the mutation patterns for one or more countries from the global framework were observed.

**Table 1:**
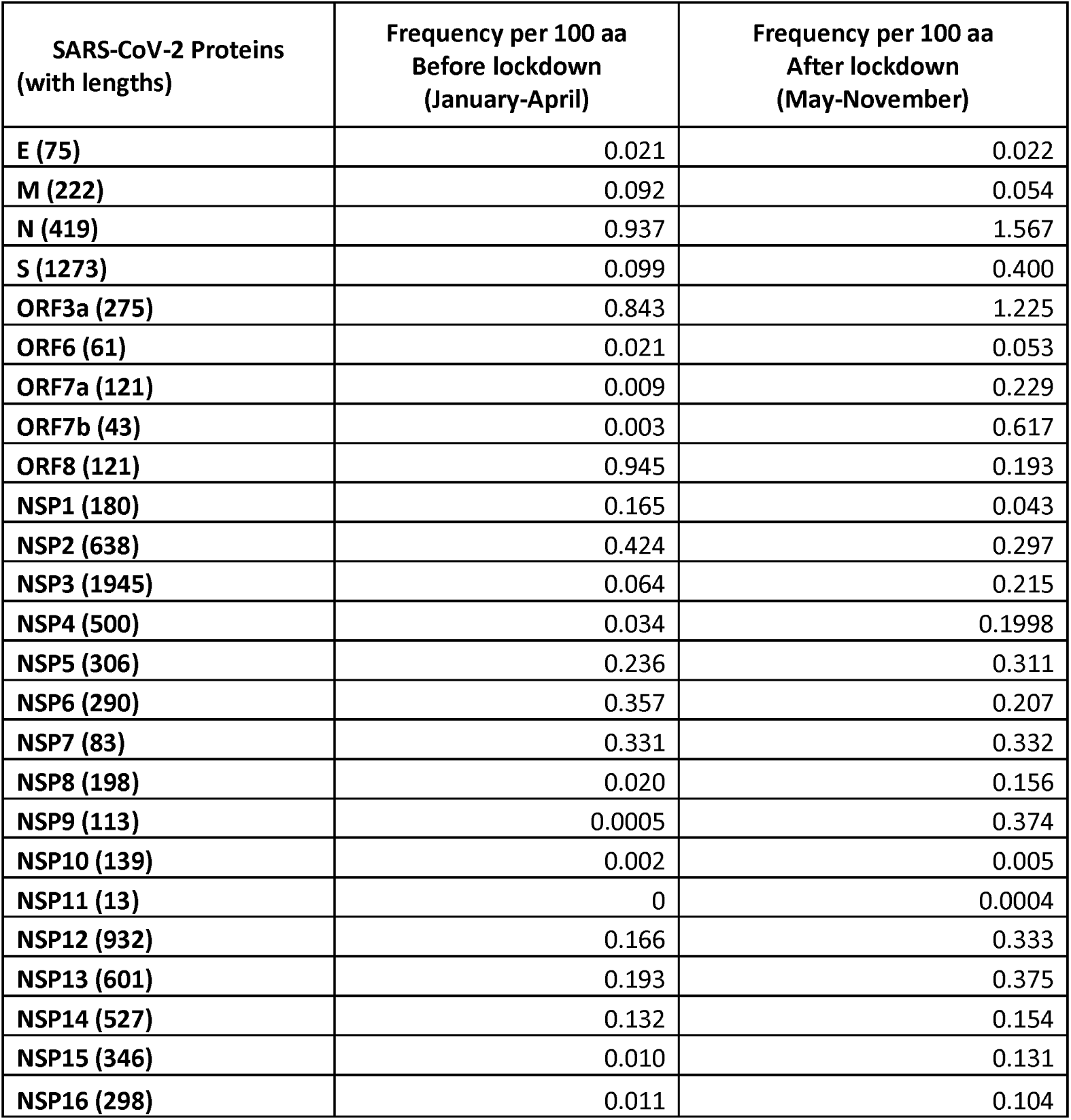
The mutation frequencies per 100 amino acids for frequently mutated residues across all countries both for pre- and post-international lockdown periods.

| **SARS-CoV-2 Proteins**<br>**(with lengths)** | **Frequency per 100 aa**<br>**Before lockdown**<br>**(January-April)** | **Frequency per 100 aa**<br>**After lockdown**<br>**(May-November)** |
| --- | --- | --- |
| **E (75)** | 0.021 | 0.022 |
| **M (222)** | 0.092 | 0.054 |
| **N (419)** | 0.937 | 1.567 |
| **S (1273)** | 0.099 | 0.400 |
| **ORF3a (275)** | 0.843 | 1.225 |
| **ORF6 (61)** | 0.021 | 0.053 |
| **ORF7a (121)** | 0.009 | 0.229 |
| **ORF7b (43)** | 0.003 | 0.617 |
| **ORF8 (121)** | 0.945 | 0.193 |
| **NSP1 (180)** | 0.165 | 0.043 |
| **NSP2 (638)** | 0.424 | 0.297 |
| **NSP3 (1945)** | 0.064 | 0.215 |
| **NSP4 (500)** | 0.034 | 0.1998 |
| **NSP5 (306)** | 0.236 | 0.311 |
| **NSP6 (290)** | 0.357 | 0.207 |
| **NSP7 (83)** | 0.331 | 0.332 |
| **NSP8 (198)** | 0.020 | 0.156 |
| **NSP9 (113)** | 0.0005 | 0.374 |
| **NSP10 (139)** | 0.002 | 0.005 |
| **NSP11 (13)** | 0 | 0.0004 |
| **NSP12 (932)** | 0.166 | 0.333 |
| **NSP13 (601)** | 0.193 | 0.375 |
| **NSP14 (527)** | 0.132 | 0.154 |
| **NSP15 (346)** | 0.010 | 0.131 |
| **NSP16 (298)** | 0.011 | 0.104 |

### 2.5 Classifying the different mutations on account of their temporal variations

The mutated residues were grouped into three classes. The first of these occurred with a frequency of 10% and higher in at least one of the countries in group A and B were considered to have originated before the stipulation of inter-country travel ban and continued to prevail, either in the previous location or in other regions around the globe. On the contrary, mutations which transpired in 10% of the strains in the early months of the pandemic but were observed to dwindle down after May were bracketed in the second class, whereas mutations that were virtually non-existent before May but emerged with higher frequencies during the ensuing months were placed in the third class. A heatmap depicting the frequencies of these mutations in their respective groups was generated using the R package ‘gplots’, where hierarchical clustering of the various substitutions was carried out using ‘Ward.D2’ method (**Figure 1)**.

### 2.6 Tracking the evolution of the mutated residues over time in a bi-monthly basis

For each of the mutations comprising the three groups, we have calculated their frequency of occurrences in every two months, from January to October 2020. Since the amount of sequencing in each country is largely inconsistent, in order to capture the mutational frequencies with a fair degree of confidence, we chose to include the total sequences for every two months for frequency calculation instead of conducting the study on a monthly basis. The frequencies were calculated only for those countries in which the mutation was observed to occur with a minimum of 10% abundance in at least one of the time-points considered.

### 2.7 Country specific co-occurring amino acid substitutions

We carried out analysis of co-occurring amino acid substitutions, considering those that have occurred with 10% or higher frequencies in each country, encompassing all viral proteins and have analysed for the residues which have co-occurred with higher abundance. This allowed us to identify the viral variants which were showing greater dominance in a certain region along with the co-occurring mutations in the respective variants. We have also discerned the substitutions which had occurred simultaneously in the countries prone to those substitutions, and have further corroborated the fact from the bi-monthly frequencies of such variants.

### 2.8 Overlapping the frequent mutations in the S-protein with predicted epitopes from external studies

A number of studies have been carried out for speedy engineering of vaccines against the SARS-CoV-2 virus. We selected eight studies which have experimentally determined potential immune-responsive epitopes owing to several viral proteins, of which we are interested especially with the epitopes delegated towards the Spike protein (Farrera-Soler, Daguer et al. 2020, Li, Lai et al. 2020, Mishra, Huang et al. 2020, Poh, Carissimo et al. 2020, Shomuradova, Vagida et al. 2020, Wang, Wu et al. 2020, Yi, Ling et al. 2020, Musico, Frigerio et al. 2021) (**Supplementary Table 5a)**. Several studies have used in-silico approaches to predict T-cell and B-cell epitopes that could potentially be employed to trigger immune responses in the host. We have selected three such independent studies which used different methodologies to predict for putative epitopes that could be implemented in designing safe and effective vaccines (Cun, Li et al. 2020, He, Huang et al. 2020, Yarmarkovich, Warrington et al. 2020) (**Supplementary Table 5b**). These potential epitopes were intersected with all frequently mutated S-protein substitutions obtained in our study. The mutations found to be harboured within one or more of the epitopic sites were observed along with the respective countries in which they were dominant. To further ascertain the progression of these mutants over time, we studied the bi-monthly frequencies of these variants.

## 3. RESULTS

### 3.1 Frequently mutated residues in SARS-CoV-2 during pre- and post-travel ban

To assess the effect of international travel restrictions in shaping the mutational profile in 35 countries, all protein sequences of SARS-CoV-2 comprising ∼2,15,000 isolates were analysed. We identified 334 amino acid substitutions that were observed in at least 2% of the total number of isolates, for at least one country in the period from January to April 2020, while 656 such frequently mutated residues were obtained in the interval from May to November 2020 **(Supplementary Tables 1a and 1b)**. Among the structural proteins, the Envelope and Membrane proteins were the most resilient to non-synonymous changes. On the other hand, the Spike and Nucleocapsid proteins displayed greater plasticity, respectively accounting for 41 and 37substitutions during the initial four months (group B, for ‘*before*’), and 80 and 69 substitutions during post travel ban (group A, for ‘*after*’). ORF3a, ORF8 and ORF7a were the most mutable in their respective phases. Expectedly, ORF1ab, which constitutes around 70% of the viral genome, represented by 16 NSPs had encountered the most number of substitutions, cumulatively equalling to 214 and 408substitutions in Group B and A respectively. Probing into the substitutions sustained by the NSPs, NSPs 2, 3 and the replication transcription complex (RTC) proteins NSP-12, 13 and 14 were the highest in the group. The summary tables containing the frequencies of each of the frequently mutated residues, as observed in 35 countries for both groups are presented in **Supplementary Tables 1a and 1b**.Overall, we found that the number of mutations in Group A had at least doubled compared to their status as observed in Group B.

### 3.2 Non-synonymous mutations per 100 amino acids in the pre- and post-lockdown

A weighted average of the frequently mutated residues per 100 amino acids (≥ 2% of all variants in a country) across all countries was calculated separately for both Group B and A **(Table 1)**. For the months preceding the travel ban restrictions, the global trend suggested that the mutation rate per 100 aa was the highest for ORF3a(0.84%)and ORF8 (0.94%)for the non-structural proteins while Nucleocapsid (0.93%)exhibited the highest rate among the structural proteins. When calculated for the following months, Nucleocapsid(1.56%), Spike proteins (0.4%), ORF3a (1.22%)and ORF7b(0.61%)were found to have the highest rates of mutations. Nucleocapsid, ORF3a and ORF7b showed the largest surge in mutation rates over the two phases, while ORF8 depicted opposite behaviour with an overall decrease in rate by 0.75 residues per 100 aa**(Table 1)**.The structural proteins E and M continued to display remarkable robustness and remained largely conserved over the entire period.

Upon examining the rate of mutations for each country**(Supplementary Table 2)**, some differences were observed. In the months leading to the lockdowns where ORF3a had the highest mutation rates in 13 countries, followed by ORF3a and Nucleocapsid in 8 countries each. However, in the period from May to November, Nucleocapsid displayed the highest mutation rates in 18 of the 35 countries, while in 7 countries ORF3a had encountered the highest number of mutations **(Supplementary Table 2)**. Country-wise mutation rates were most stark in case of ORF8, which had remained immune to changes in 21 countries while effecting fairly high rates of substitutions in the remaining countries. Apart from these, ORF7a in South Korea, ORF7b in UK and Latvia, and ORF8 in Kenya and South Africa showed the highest rate of mutations.

### 3.3 Mutations that have occurred with relatively high frequencies (≥10%) in one or both groups under study

#### 3.3.1 Frequently mutated residues during pre- and post-travel ban

We next determined the mutations with at least 10% frequency in both the phases under investigation. This comprised a total of 29 mutations as presented in the heatmap (**Figure 1a)**.Among these, the D614G substitution in the Spike glycoprotein was seen to have reached stability among the entire global population, with frequencies touching 100% in most countries. The R203K and G204R mutations in the Nucleocapsid have increased in most of the countries, though not as sharply as the case with the Spike protein. These mutations were observed to be highest in the continents of North America, South America, Asia and Australia, while in many of the European countries the mutational rates in these two residues of the N protein have either remained the same or have decreased variably. Among other notable substitutions, Q57H of ORF3a shows conflicting trends across the countries, having decreased in several North and South American countries, while gaining in particular countries in the European conglomerate. Other notable mutations in the common set include N_S194L, which has increased in all the occurring countries, being particularly high in Mexico. On the other hand, NSP12_L323P has significantly reduced in most of the countries since May, with the exception being Singapore where it remained stable. Another instance of a gradually decreasing mutation is NSP2_T85I, where we can see the mutations reducing significantly with exceptions being Canada, Germany, Ireland, Kenya. Surprisingly, South Korea shows a large increase in this variant. In this group, we also noted several mutations to endure over the considered time period, albeit in one or only few selected countries. These include the L71F in the NSP7,S1176Fin Spike protein and I33T in ORF6, discerned only in Brazil, the substitution A994D in the non-structural protein NSP3 observed solely in India. Additionally, the ORF8 substitution S24L in USA, Spike protein variation D936Y in Sweden, NSP6_M86I and N_S202N substitutions in UAE and Kenya, NSP3_I1683T substitution in Poland, Czech Republic, Denmark and Ireland were among those that were exclusive to the specified countries.

#### 3.3.2 Mutations that have declined and/or eliminated after international travel ban

Moving on to the mutations that were observed in the period before inter-country travel ban, we notice a distinct cluster comprising mutations in N and M proteins, ORF3a and NSPs 2,3, and 4 **(Figure 1b)**. In the subsequent months, these variations were seen to have substantially decreased, with most of them being nearly wiped out. It was also interesting to observe substitutions P344S in Nucleocapsid (UAE), L781F in NSP3 (South Korea), G172C in ORF3a (Bangladesh), E120K in NSP3 (Bangladesh), S183Y in Nucleocapsid (New Zealand), L275F in ORF3a (Ireland), Y28H in Spike (UAE) were prevalent in the Group B before May while being significantly reduced in Group A.

### 3.3 Mutations that have newly emerged or increased sharply post-travel ban

We next surveyed the mutations that were virtually non-existent before May, but gained prominence in specific locations post travel ban **(Figure 1c)**. Interestingly, we noticed that the European countries formed a separate cluster comprised of4 mutations in the S protein, 3 each in Nucleocapsid and ORF3a, and others encompassing NSP4, NSP6, NSP7, NSP9, NSP12, NSP13 and NSP16. These were clearly not pervasive before May and had come into effect in the wake of the travel bans. Apart from these, other noteworthy mutations that originated in specific countries post-lockdown were found in Nucleocapsid and NSPs3, 5 and 12 in Japan, three mutations comprising the S protein, NSP3 and NSP15 in China. Additionally, substitutions exclusive to South Korea among NSP7 and NSP16, mutations in S and NSP3 specific to Czech Republic, two mutations in ORF7a and NSP3 prevalent only in Kenya, and two variations within NSP3 and Sin Canada were among the ones that were absent in the pre-lockdown era.

### 3.4 Temporal survey of the mutated variants on a bi-monthly basis

Upon identifying mutations specific to certain countries, we wanted to see whether these variations had come into being or declined sharply at a distinct time point. Estimation of bi-monthly frequencies for the corresponding countries (**Supplementary Table3)** allowed us to perceive three distinct trends with which the mutations developed in the course of time. The substitution S_D614G was seen to have attained close to 100% concordance in most countries by October, with North American and European countries gaining the mutation at a faster rate compared to Middle East and Asian countries (**Figure 2a-c)**. In contrast, the co-occurring substitutions R203K and G204R in Nucleocapsid showed more diverse variations **(Figure 2d-f)**. In most of the countries, these substitutions showed greater dominance over the wild type, with countries like Australia, Mexico, Russia, Japan, Bangladesh and Latvia, where the corresponding frequencies rose to ∼90% and higher. However, in several European countries namely Belgium, Denmark, France, Germany, Ireland, Netherlands, Norway, Spain, Switzerland and UK, and also in USA and Brazil, this variant has not gained a firm footing over its wild-type counterpart, remaining more or less constant over the specified time period or plummeting after brief ascension. ORF3a substitution Q57H continued to reign disproportionately in different countries, where it can be seen to have attained a semblance of stability in USA, Canada, Mexico and Sweden **(Figure 2g)**, while low abundance or plunging rates were observed in Czech Republic, Germany and Australia among others **(Figure 2h)**. Contrarily, Belgium, France, Switzerland and China were among the nations where this variant was seen to be on the ascension after a brief hiatus around July-August **(Figure 2i)**.

**Figure 2:**
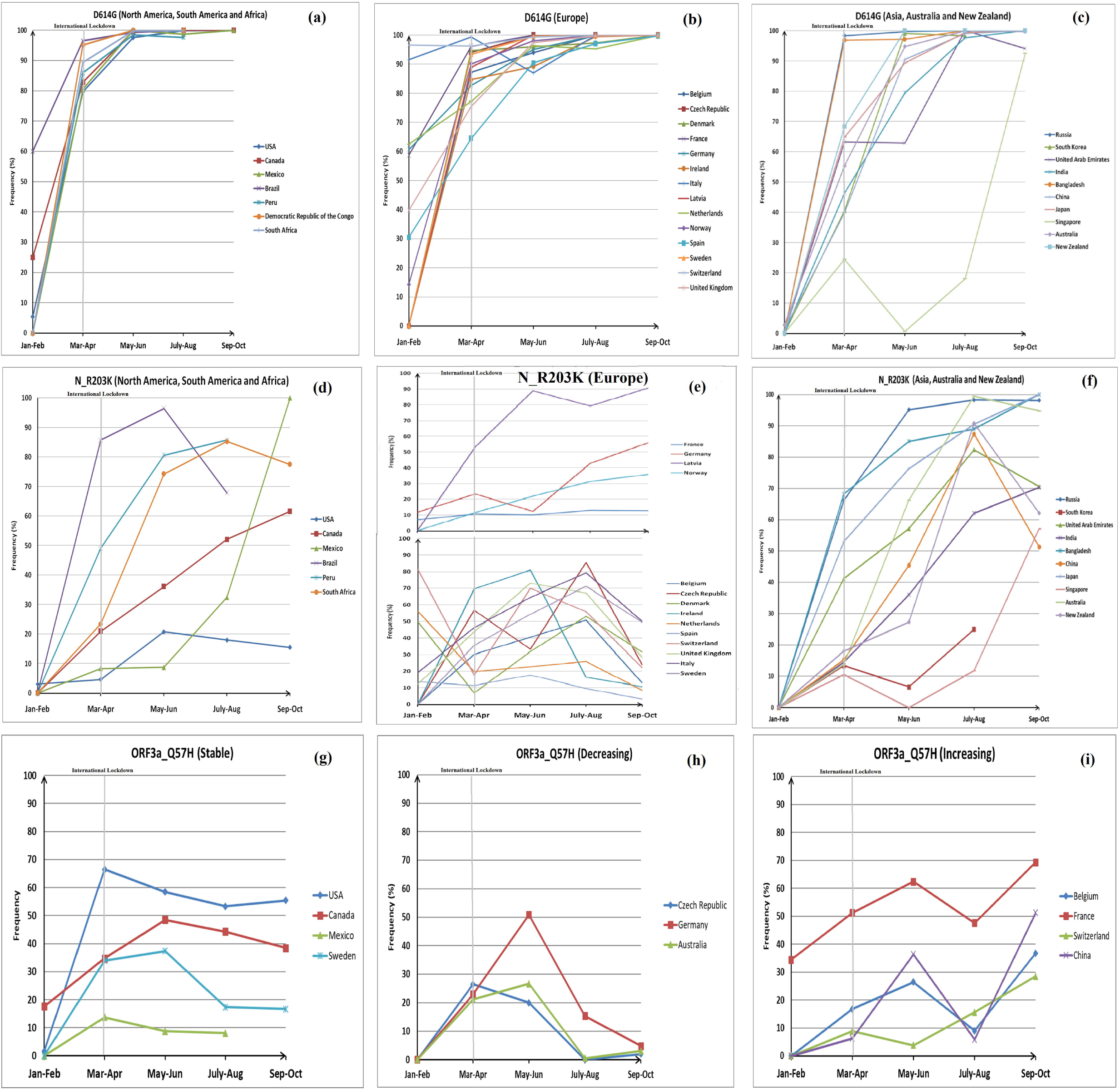
Bi-monthly frequency plots of substitutions **(a, b and c)** D614G in Spike protein, (**d, e and f)** R203K in Nucleocapsid and **(g, h and i)** Q57H in ORF3a both before and after international lockdowns. S_D614G **(a, b and c)** and N_R203K **(d, e and f)** plots have been segregated according to geographical locations, while the three distinct trends of mutation of ORF3a_Q57H **(g, h and i)** have been shown as separate plots.

There were instances of mutated variants that had gained a brief upsurge pre-travel-ban but were seen to diminish in stature in the ongoing months. D936Y and I119V substitutions in S protein specific to Sweden and Peru showed dwindling characteristics **(Figure 3a)**. A similar trend was observed for S197L substitutions in Nucleocapsid in Spain and Australia, which were coincident with F308Y substitution in NSP4 in the same countries **(Figure 3b)**.Similarly,N_P13L and NSP3_T1198K variants that were appreciably high in India and Singapore before May, sharply reduced concurrently in the later periods**(Figure 3c)**. Moreover, S183Y, Q240K and P344S in Nucleocapsid, characteristic of New Zealand, Ireland and UAE had taken a much sharper nosedive **(Figure 3d)**. NSP2_T85I with the exception of USA had peaked in the interval of March-April before following a downward trajectory **(Figure 3e)**. Among other noteworthy mutations that were on the descent after a brief period of prevalence are NSP12_L323P and ORF3a_G251V as seen in myriad nations (**Figure 3f, g**).

**Figure 3:**
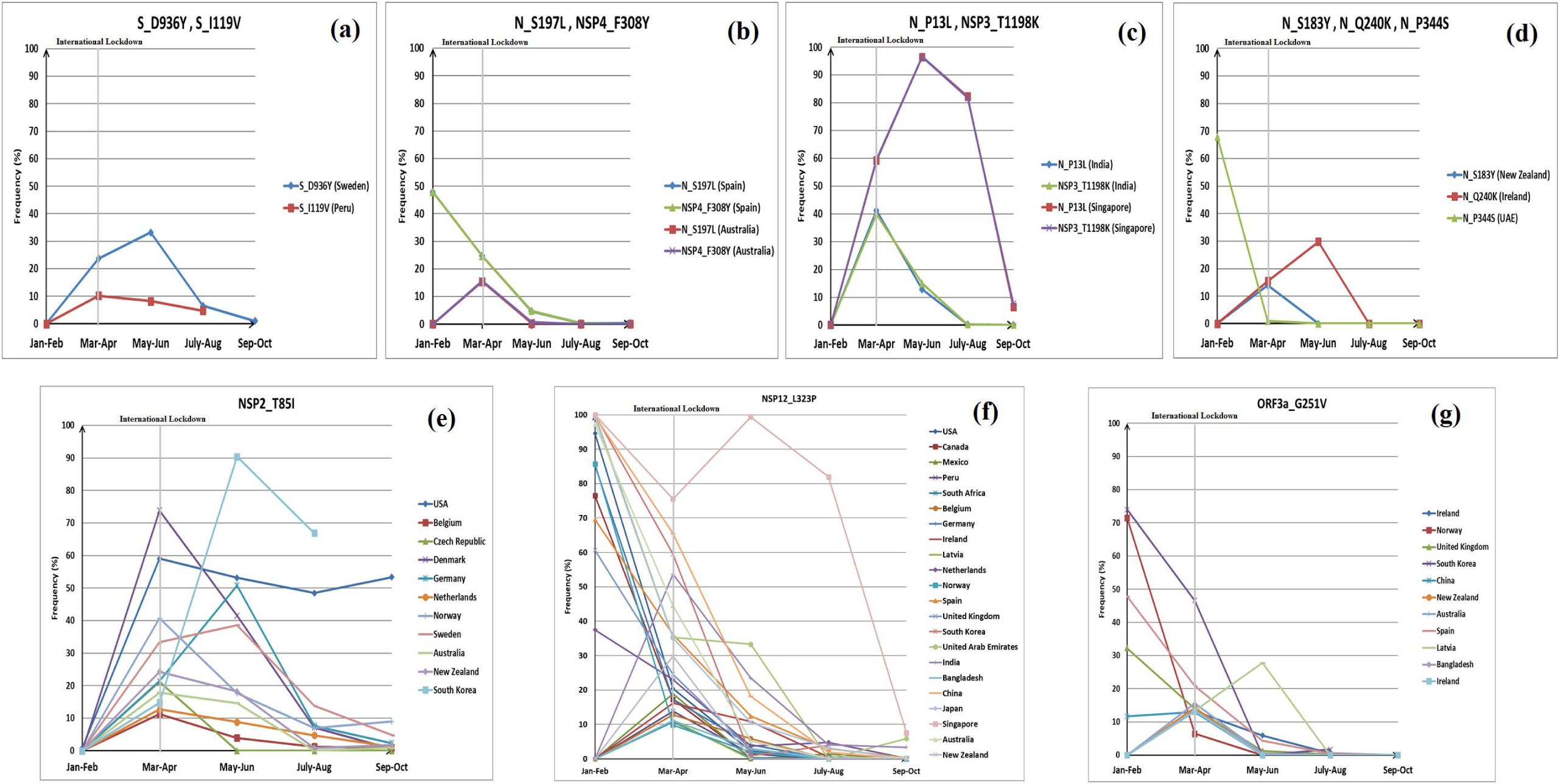
Bi-monthly plots of selected mutations seen to prevail pre-lockdown but have sharply reduced in abundance in the following months. Various mutations have been shown in representative countries corresponding to the **(a-d)** S and N proteins, **(e)** NSP2, **(f)** NSP12 and **(g)** ORF3a.

Several new mutations were found to have emanated after May and were seen to be on the rise in specific countries. Our interest was specifically arrested by two substitutions, A222V (**Figure 4a**) and S477N **(Figure 4b**) in S protein that were seen to emerge after July and progress rapidly thereafter, the only exception being Norway **(Figure 4a)**. These mutations were exclusively obtained in high percentages largely among the European nations, with the exception of the 477N variant also being widespread and on the rise in Australia **(Figure 4b)**. Among other variants, Spike protein mutations on the ascension post-travel-ban stage included S98F exclusive to Belgium and Netherlands, L18F in the UK and V1122L in Sweden **(Figure 4c)**. N-protein variantsR40C specific to Latvia and P151L in Japan were seen to have reached 50% abundance by October. Among the NSPs, significant mutations that were on the ascendency include NSP2 substitution I120F exclusive to Australia and Bangladesh, coincidentNSP3 substitutions Q203R and S284G in Latvia andI441V in Belgium and S543P in Japan, NSP5_L89F and NSP5_P108S in USA and Japan respectively, P59T variant of NSP10 occurring in Latvia, A423V and V720I mutations of NSP12 in Japan and Czech Republic respectively **(Figure 4d)**.

**Figure 4:**
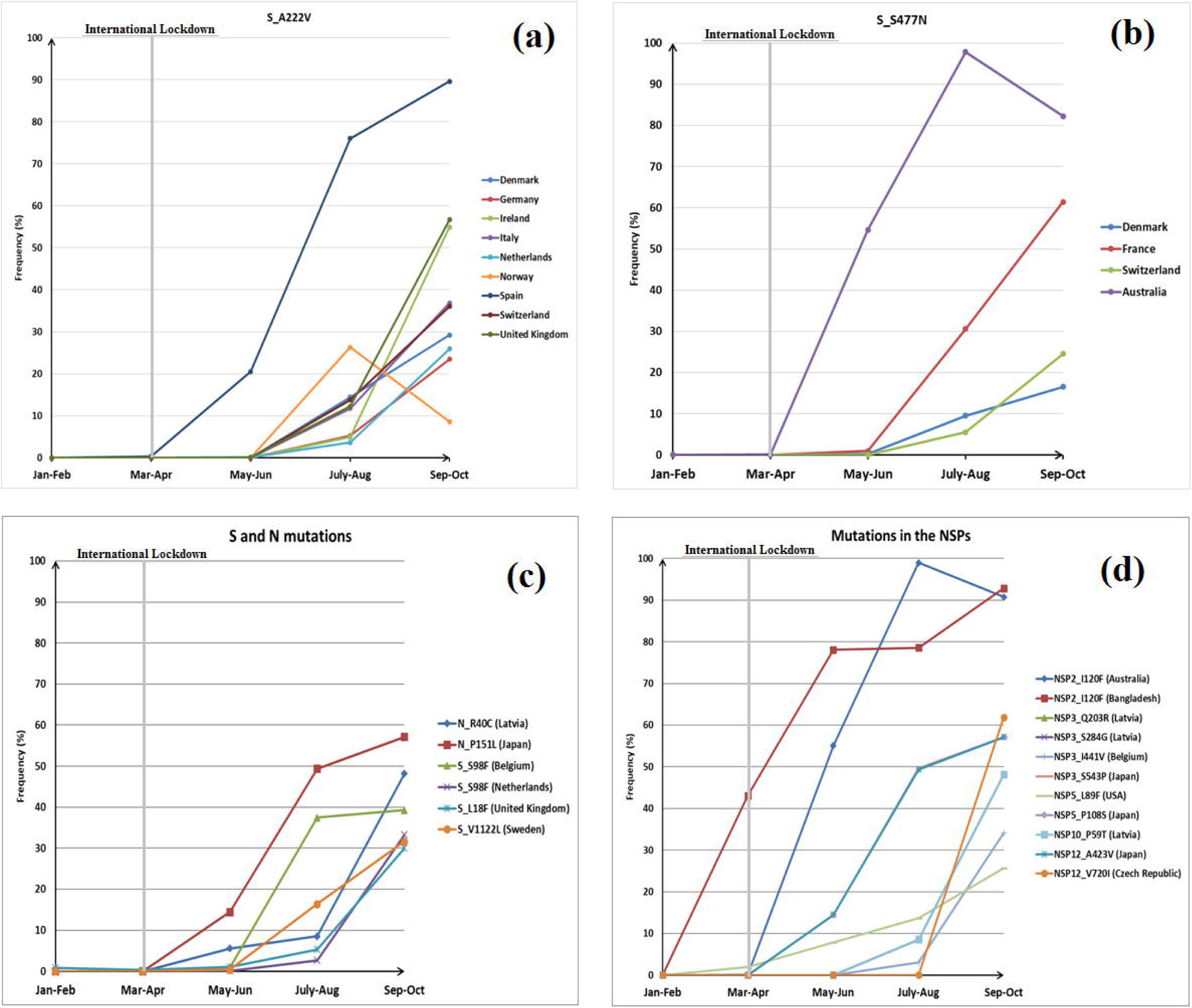
Bi-monthly plots depicting the mutations that had emerged in specific countries after imposition of travel ban. Spike protein mutations **(a)** A222V and **(b)** S477N were found to have increased specific to European countries. Remaining mutations in the **(c)** Spike and Nucleocapsid proteins, and **(d)** non-structural proteins corresponding to distinct countries.

### 3.5 Mutations that have co-occurred with identical or nearly identical frequencies

Now that we had identified prominent mutations in diverse SARS-CoV-2 proteins, we wanted to find the substitutions which have co-occurred in different geographical locations. We have considered variants with a prevalence of at least 10% for a particular country to be the dominant variant. These prevailing variants arising from the merger of various proteins along with the corresponding frequencies are presented in **Supplementary Table 4**. We have noticed that with the exception of China, all the substitutions have occurred in the background of the Spike protein D614G variant, which in all likelihoods preceded the inception of the other mutations. Apart from the known co-occurring mutations in the Nucleocapsid protein at 203^rd^ and 204^th^ residues, we noted several other mutations that have appeared concurrently as also verified by having similar frequencies in one or more countries. Spike protein variant A222V went hand-in-hand with the A220V variant in the Nucleocapsid, seen exclusively in the European nations. F308Y substitution in NSP4 co-occurred with S197L variation in the Nucleocapsid, as observed in Spain and Australia. However, these resulting variants did not prosper and were seen to be obliterated over time. Substitutions G15S and T428I in NSP5 and NSP3 respectively were observed in Peru, Canada, South Africa and New Zealand with appreciable frequencies. These mutations were gained after May but were seen to be decreasing as of October. Substitutions M234I and A376T in the Nucleocapsid, M324I in NSP4, A185S and V776L in NSP12 alongside K218R and E261D in NSP13 were seen to have arisen with identical measures after June and have reached appreciable levels in Belgium, Denmark, France and Switzerland.NSP13 also encountered simultaneous substitutions in 504^th^ and 541^st^ residues as seen in USA. ORF3a had similarly manifested corresponding mutations in 172^nd^ and 202^nd^ residues observed in Belgium and Netherlands from the month of July (**Supplementary Table 3**).

### 3.6 Mapping the frequently mutated residues with Spike protein epitopes

We have mapped the frequently mutated residues observed in the post-lockdown period with S protein epitopes obtained from 11 studies (**Figure5, Supplementary Table 5a,b)**. Among the studies concerned with experimentally determined epitopes, one study (Farrera-Soler, Daguer et al. 2020) escaped convergence with any frequently mutated residues. The remaining reports contained at least one site which overlapped with the observed mutations in the Spike protein. The list of experimentally predicted epitopes with the corresponding mutations nestled within them along with the countries are provided in **Supplementary Table 6**. Bi-monthly frequencies were calculated for each of these mutations (**Supplementary Table 7**) to trace the course of their advancement through time, and determine those mutants, which might seem innocuous in the early stages but could pose a threat to vaccine potency if left unchecked.

**Figure 5:**
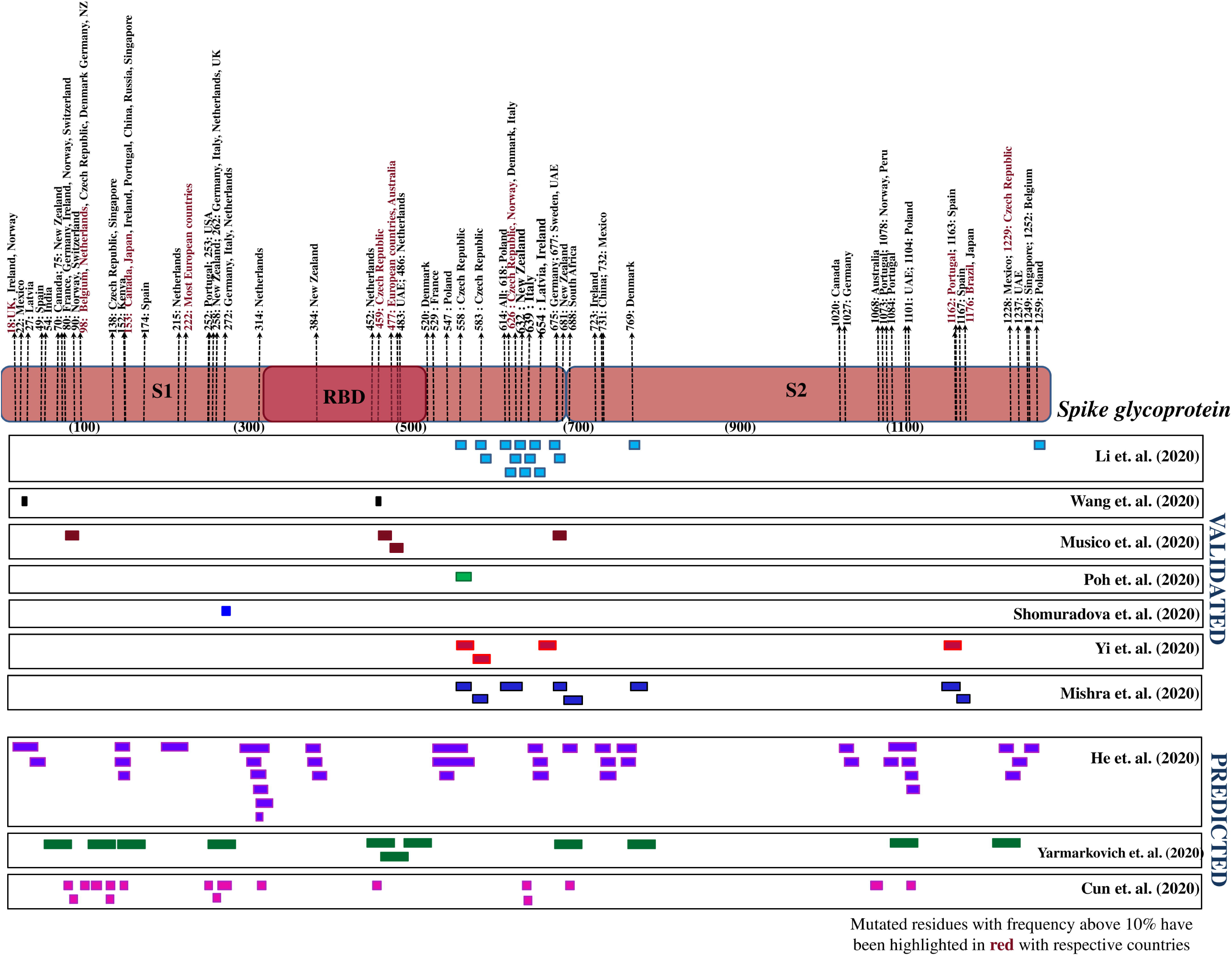
Illustration of the predicted Spike protein epitopes which have overlapped with one or more frequently mutated Spike protein residues (Cun, Li et al. 2020, He, Huang et al. 2020, Li, Lai et al. 2020, Mishra, Huang et al. 2020, Poh, Carissimo et al. 2020, Shomuradova, Vagida et al. 2020, Wang, Wu et al. 2020, Yarmarkovich, Warrington et al. 2020, Yi, Ling et al. 2020, Musico, Frigerio et al. 2021). The countries in which they were found to be prevalent post lockdown have been mentioned alongside. The residues with frequencies greater than 10% have been highlighted along with the corresponding countries.

Most of the studies predicted epitopes within 500 to 700 amino acids positions of S-protein (**Figure 5)**. This region contained perceived mutational changes, chief among them are substitutions at positions 614 and 626 of S protein. The A626S variant exhibited contrasting trends in Czech Republic as opposed to Norway, Italy and Denmark. In both cases this mutant came into existence July onwards, but diminished sharply after peaking in Czech Republic, while was on the rise in the remaining countries. Other noteworthy mutations which may compromise vaccine effectiveness include T632I (New Zealand), G639S (Italy), E654A (Ireland), Q675H (Germany), Q677H (Sweden, UAE) and P681H (New Zealand), all of which were seen to increase in the months after travel ban albeit at varying rates. Moreover, theV1176Fvariant in Brazil warrants immediate attention as these were seen to accumulate substantially in the span of ten months. Mutations D1163Y and G1167V, both identified in Spain should not be discounted as these were also on the ascendancy as of October, while being present within a putative epitope as outlined by at least 2 studies. Another variant P1162S, shared within epitopes predicted by these studies was observed with a frequency greater than 10% in Portugal around the time of travel ban but lack of sufficient sequencing data in the latter months deprives us to get information of its evolution. Another substitution S459Y soaring over 10% frequency in Czech Republic was also overlapped with predicted epitopes by two studies. However, this variant were seen to subside in abundance by October in the respective countries. A solitary epitope contained P272L substitution which was discerned to rapidly gain grounds in many European nations. Similarly, S477N variant present in one epitopic site was already seen to be abundant in Australia and numerous European nations. Other epitope containing mutations that had an upwards curvature included V483A in UAE, seen only after September, F486L in Netherlands and V90F in Norway and Switzerland seen from July onwards.

The in-silico epitope predictions likewise mapped with several substitutions, the details of which are presented in **Supplementary Table 8**. These included substitutions L18F, seen to be increasing in UK, A222V rampaging among the European population and M1229I specific to Czech Republic. Among other substitutions, S98F variant as seen previously, exhibited an upwards trajectory, more noticeable in Belgium and Netherlands, M153T and S459Y variants were uncovered in Japan and Czech Republic respectively, while S477N was a major mutant that had disseminated across the European countries.

## 4. DISCUSSIONS

We carried out a thorough country-wise characterization of the mutational landscape of SARS-CoV-2. Upon comparing the mutational profile of individual countries before and after the cross-country travel bans allowed us to perceive the evolutionary trajectories of pre-existing as well as newly erupted variants. The spike protein D614G, which had been characterized to have superior transmission rates and stabilities compared to the D variant(Hou, Wang et al. 2007, Korber, Fischer et al. 2020, Laha, Chakraborty et al. 2020), was found to have reached 100% confluence in most countries. Other previously reported variations in the N, ORF3a, ORF8 were also seen to prevail in multiple nations with varying degrees, which could be attributed to the diverse ethnic backgrounds or environmental conditions of the different regions. Several mutations that had materialized before April in certain countries, were observed to have been wiped out from the endemic populations after a brief period of dominance. One of the explanations of this phenomenon could be that the resulting variations greatly diminished the stability or transmissibility of the new SARS-CoV-2 variants, which led to their gradual elimination. Another hypothesis that could be attributed to this is that the new mutations were highly lethal in nature, and the infected individuals upon contracting these variants would consequently be placed under stringent quarantine, thereby preventing the propagation of such variants in larger numbers. Other factors such as the frequency of international travel to and from these countries during the concerned period, as well as the extent of lockdowns in these countries could have a say in the final outcome of these variants. One could try to draw a correlation between the period of prevalence of these mutations and the mortality rates of the associated countries, in order to corroborate this speculation.

We were essentially interested to study whether the travel-ban could lead to the appearances of country-specific mutations. Several new strains have been found to originate after May, which were seen to increase with the passage of months. Particular attention should be drawn towards the European countries which have been the epicentre for few major variants especially the spike protein variants A222V and S477N and the N protein substitutions A220V, M234I and A376T. From the bi-monthly frequency data, we could visibly track the upward progress of these variants, and with recent relaxations in travel bans, these strains could in short time pervade the globe.

Extensive efforts to design safe and efficacious vaccines were already in full swing since the early days of the pandemic. Structural proteins, especially the Spike glycoprotein are the preferred targets for vaccines as they are directly involved with fusion with and entry into the host cells. So far most of the mRNA-vaccines have been designed to target the receptor-binding domain (RBD) of the Spike glycoprotein, which has shown to elicit immune response and synthesize antibodies when experimented in murine cell models and the sera of humans who had contracted the disease. However, other proteins like Nucleocapsid, ORF6, ORF8, NSP3 can also be viewed as potential candidates when designing multi-epitope vaccines against the virus(Chukwudozie, Chukwuanukwu et al. 2020, Dong, Dai et al. 2020, Dutta, Mazumdar et al. 2020, Mandala, McKay et al. 2020). As of December 2020, as many as 55 vaccines have entered the foray of clinical trials with many more yet in their preliminary stages. Among the frontrunners that have gained approval include the mRNA-based vaccines named BNT162b2 (Polack, Thomas et al. 2020)and mRNA-1273(Baden, El Sahly et al. 2020)manufactured by ModernaTX Inc.; CoronaVac(Zhang, Zeng et al. 2020), which utilize an inactivated form of the virus; Sputnik V(Balakrishnan 2020, Burki 2020). However, since Spike proteins only comprise a small fraction of the viral proteome, epitopes focusing only on this singular protein may not be as effective in eliciting a strong immune response compared to a mutli-epitope vaccine designed to target multiple viral proteins. Several of the SARS-CoV-2 proteins including Spike, Nucleocapsid, NSP5 (Mpro) and NSP12 (RdRp) have been successfully crystallized (Kirchdoerfer and Ward 2019, Jin, Du et al. 2020, Kang, Yang et al. 2020, Lan, Ge et al. 2020, Lee, Worrall et al. 2020), which can be utilized in engineering the therapeutic agents. Several bioinformatic approaches and in-silico tools have been developed which have predicted peptides that could be used as epitopes, in inducing immune response against the virus. However, at the time of contriving these predictions, some of these novel variants that have been uncovered only after a certain time point would not have been taken into account. We have intersected the predicted epitopes from a total of 11 independent studies, 8 of them corroborated by experimental results. A handful of them coincided with highly mutable sites while several mutations originating in the months following lockdown and were seen to be on the rise thereafter. Among these the residue at positions 626 and 477 need close supervision while selecting epitopes as vaccine candidates.

The vaccines which recruit the whole virus in inactivated forms such as PicoVacc(Burki 2020)and BBIBP-Corv (Wang, Zhang et al. 2020, Isakova-Sivak and Rudenko 2021), can code for the structural as well as accessory viral proteins in the host cells, which are desirable as they present multiple epitopic sites for antibody production. Though, the M and E-proteins are largely conserved, they have been found to display poor immunogenicity (Du, He et al. 2008). The Nucleocapsid, on the other hand, though found to be highly immunogenic failed to provide adequate protection against SARS-CoV-2 when tested in mice models (Sun, Zhuang et al. 2020). We have obtained several mutations within the Nucleocapsid proteins in diverse countries, chief among them being the modifications at positions 13, 220, 234 and 376 which should not be disregarded. A point that needs to be highlighted here is that the antibodies induced by the vaccines are polyclonal in nature. Therefore, even if one of the epitopic sites of the antibodies ate rendered ineffective owing to mutational changes, the other veritable epitopic sites should come into play in combating the virus instead of the antibodies being entirely dispensable. Only the degree of efficacy of the vaccines may vary among the nations. Moreover, new viral strains are continually cropping up in different countries, with a novel, highly contagious strain being recently reported in the United Kingdom(Wise 2020). The N501Y mutation, a key substitution within the UK strain, was inspected for in our study in the months of September and October, where we have found it to be present in only three countries, with highest among them being UK (0.5%), followed by South Africa (0.22%) and USA (0.03%). Therefore, it is comprehensible that the meteoric rise of this variant emanated around the month of December. Analysing for any other emerging mutations with frequencies above 2% in the Spike protein, in the peripheral months of September and October as per out study, we have detected several new substitutions among various countries, with the variants L179F and Q913H in Japan and F486L in Netherlands to have frequencies above 10% (**Supplementary Table 9**). These mutants had not been observed with noteworthy frequencies in the preceding months, but only came into prominence towards the later period. As we cannot state with any degree of certainty the evolutionary course these variants will follow, countries should actively monitor these sites in the coming months. If any of these mutations are inadvertently incorporated among the epitopes of future vaccines, they may render them partly impotent in countries where those variants are thriving. Now that there have been considerable relaxations in inter-country migrations, other nations should also follow up the status of these mutations to confirm the fidelities of the associated amino acid residues. We conclude with the hope that the worst days of the pandemic are behind us, and await the timely disbursement of safe and effective vaccines while continuing to follow adequate social distancing norms around the world.

## Data Availability

All relevant data associated with the manuscript are provided as supplementary materials.

## Acknowledgement

This work was supported by the funding of Indian Statistical Institute, Kolkata, and the CSIR for providing the fellowship to SL.

## Author contributions

RC conceptualize the study. SL performed the data analysis, SL and RC wrote the manuscript.

